# Pain-Reporting Variability and Reliability as Predictors of Placebo Effects: A Cross-Sectional Experimental Pain Study

**DOI:** 10.64898/2026.07.31.26359372

**Authors:** James A. Cottam, Yang Wang, Titilola Akintola, John Farrar, Chixiang Chen, Patrick McArdle, Seth A. Ament, Philip Corlett, Susan G. Dorsey, Roi Treister, Luana Colloca

## Abstract

Placebo analgesia varies substantially across individuals, yet the sources of this heterogeneity remain incompletely understood. Pain-reporting variability may represent an underrecognized predictor of placebo responsiveness. This study examined whether variability and reliability of pain reporting predict placebo analgesia in an experimental pain setting.

Eight hundred and three participants (401 individuals with temporomandibular disorder and 400 healthy controls) completed a standardized thermal pain paradigm involving calibration, placebo conditioning, and testing phases. We obtained repeated heat temperatures (four) during thermal calibration and pain ratings during conditioning test (24 trials). We used these measurements to quantify within-person pain-reporting variability using standard deviation (SD) and coefficient of variation (CoV), and reliability using intraclass correlation coefficients (ICC). Placebo analgesia was calculated as the difference in pain ratings between control and placebo cue trials during testing. Regression models examined associations between reporter characteristics and placebo analgesia controlling for age, sex, race and experimenters.

Variability of thermal pain responses during calibration did not predict placebo analgesia. Greater pain-reporting variability during conditioning was associated with reduced placebo analgesia (higher SD and CoV), and this effect was mediated by slower acquisition of the cue– pain contingency. Conditioning-phase reliability was not associated with placebo analgesia, whereas three clusters of learning profiles predicted placebo effects. Thus, individual differences in pain-reporting variability during conditioning, rather than baseline sensory variability, contribute to heterogeneity in placebo analgesia. These findings suggest that variability during acquisition processing matters more than general sensory variability influencing the magnitude of placebo effects.

## Introduction

Placebo analgesia is a psychobiological phenomenon that contributes substantially to pain outcomes in both experimental and clinical settings [10; 14; 39]. However, individuals vary considerably in the magnitude of placebo responses, and the factors underlying this heterogeneity remain incompletely understood [38; 43]. Although expectancy, learning mechanisms, and contextual factors are well-established determinants of placebo analgesia [7], identifying individual characteristics that predict placebo responsiveness remains important for understanding placebo mechanisms and improving analgesic research.

One potential source of variability lies in the way individuals perceive and report pain [28]. Pain ratings are inherently subjective and fluctuate across repeated assessments because of sensory, cognitive, affective, and contextual influences [26; 33]. Consequently, repeated pain assessments capture dynamic characteristics beyond mean pain intensity. Intra-individual variability can be quantified using several complementary indices, including standard deviation, autocorrelation, mean square successive differences, and probability of acute change [25; 26].

Evidence suggests that within-subject pain-report variability may influence placebo responsiveness [37], although findings have been inconsistent. Most studies have focused on variability in spontaneous clinical pain recorded before treatment using daily pain diaries or ecological momentary assessment [13; 17]. In several chronic pain trials, greater baseline pain variability was associated with larger placebo responses without affecting active treatment response, suggesting that unstable pain reporting may reduce assay sensitivity and contribute to placebo-related improvement. These findings prompted consideration of baseline pain variability in clinical trial design [16]. However, other studies have failed to replicate these associations [19], indicating that the predictive value of baseline pain variability remains uncertain and may depend on the pain condition, study design, or how variability is measured.

Previous studies have primarily examined variability in spontaneous clinical pain using daily diaries or ecological momentary assessment [13; 17]. More recently, experimentally evoked pain variability, particularly using the Focused Analgesia Selection Test (FAST), has also been associated with placebo responsiveness, although findings remain limited and inconsistent [31; 36; 37]. Whether variability during standard quantitative sensory testing and placebo conditioning similarly predicts placebo analgesia remains unknown. Experimental pain precisely controls stimulus intensity [2; 4], allowing pain-report variability to be examined independently of fluctuations in nociceptive input. In addition, the reliability of pain reporting has received comparatively little attention.

Importantly, the interpretation of pain variability differs across experimental, acute, and chronic pain settings. In chronic pain trials, pretreatment variability reflects fluctuations in ongoing illness-related pain before therapy begins [13; 17]. In acute pain trials, pretreatment variability is difficult to characterize because treatment is initiated shortly after pain onset. Moreover, the consistency with which individuals rate comparable sensory experiences across repeated assessments [20], has been overlooked.

Building on previous work [8; 41], we examined whether variability and reliability of experimentally evoked pain during thermal calibration and placebo conditioning predict placebo analgesia in healthy individuals and participants with temporomandibular disorders (TMD). Variability reflects fluctuations in pain ratings across repeated assessments, whereas reliability reflects the consistency of pain ratings across repeated exposures to comparable stimuli. Examining both measures may provide complementary insights into individual differences in pain processing and placebo learning.

## Methods

This report is based on retrospective analysis of data collected between August 2016 and February 2020 at the University of Maryland School of Nursing as part of an experimental placebo study described previously [8].

### Participants

Participants aged 18–65 years were recruited. A total of 834 individuals were screened, including 421 individuals with TMD (102 men, 319 women) and 413 healthy controls (168 men, 245 women, see CONSORT file). Thirty-one individuals were excluded during screening because of major medical or psychiatric conditions, lifetime dependence on alcohol or recreational drugs, pregnancy, or use of antipsychotic medications. One participant withdrew during screening, and one was lost to follow-up, resulting in a final sample of 401 participants with TMD and 400 healthy controls.

The study was approved by the University of Maryland Baltimore Institutional Review Board (Prot# HP_00068315), and all participants provided written informed consent before study participation, compensated for their participation ($50), and debriefed at the end of the study for the deceptive component of the placebo induction (see below).

TMD diagnoses were confirmed by an orofacial pain specialist at the Brotman Facial Pain Clinic, University of Maryland School of Dentistry. Eligible participants met the Axis I Diagnostic Criteria for Temporomandibular Disorders (DC/TMD) and reported pain for at least three months [30; 42].

Healthy controls were required to have no history of chronic pain or TMD and were frequency matched to the TMD cohort by age and sex. Eligibility was confirmed during an in-person evaluation by trained study personnel. General exclusion criteria for both groups included degenerative neuromuscular, cardiovascular, neurological, renal, hepatic, or pulmonary disease; active cancer or cancer within the previous three years; uncorrected hearing impairment or color blindness; pregnancy or breastfeeding; and severe psychiatric disorders (e.g., schizophrenia, bipolar disorder, major depression, obsessive-compulsive disorder, or dementia) or lifetime dependence on alcohol or recreational drugs.

### Experimental Procedure

All experimental sessions were conducted at University of Maryland School of Nursing, Baltimore. Following informed consent, participants completed a standardized thermal calibration procedure to characterize individual heat sensitivity and determine the stimulus intensities used during the placebo procedure.

### Heat pain calibration

Briefly, thermal stimuli were delivered to the nondominant volar forearm using a computerized contact heat stimulator (30X30 mm, Medoc Pathway System, Medoc Advanced Medical Systems, Ramat Yishai, Israel). Pain intensity was rated on a 10 Numerical Rating Scale (NRS), where 0 indicated “no pain” and 10 indicated “maximum tolerable pain”.

To characterize individual thermal sensitivity, participants completed a standardized heat pain calibration procedure. Thermal stimuli were delivered in ascending order from warmth detection threshold (4 trials) to pain threshold (4 trials), moderate pain intensity (4 trials) and finally heat pain tolerance (4 trials). The ramp up and ramp down rate for warmth threshold and pain threshold were 0.8 Celsius/second, and 1 Celsius/second, respectively. The ramp up and down rate for moderate pain and pain tolerance was 1 Celsius/second and 8 Celsius/second, respectively. *Warmth detection threshold* was defined as the lowest temperature at which participants first perceived warmth, with a pain rating which should remain at 0/10 on the NRS. *Pain threshold* was defined as the lowest temperature at which the sensation was perceived as painful, corresponding to a score of 1/10 on the NRS. *Moderate pain intensity* was defined as the temperature producing a pain rating of approximately 6/10 on the NRS, and *heat pain tolerance* was defined as the highest temperature participants were willing to tolerate before terminating the stimulus, equaling a score of 10/10 on the NRS. Each thermal intensity was presented four times, allowing repeated pain intensities for each modality to be obtained. These repeated pain intensity assessments were used to quantify individual differences in pain-reporting variability and reliability (Fig. 2A). The temperature corresponding to moderate pain intensity (approximately 6/10 NRS) was selected as the individualized painful stimulus for the subsequent placebo conditioning and testing phases.

**Figure 1:**
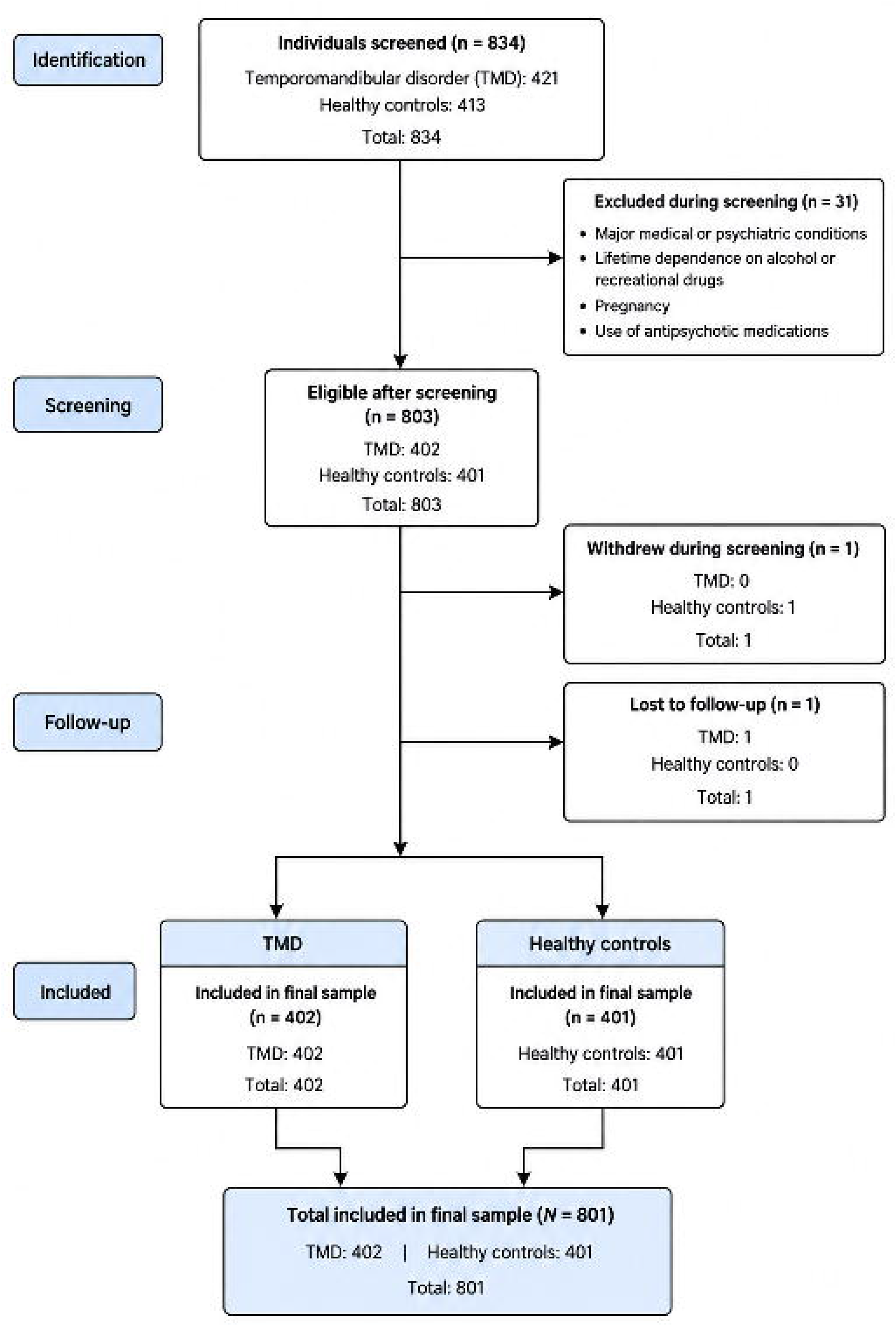
Consort flow-chart.

**Figure 2:**
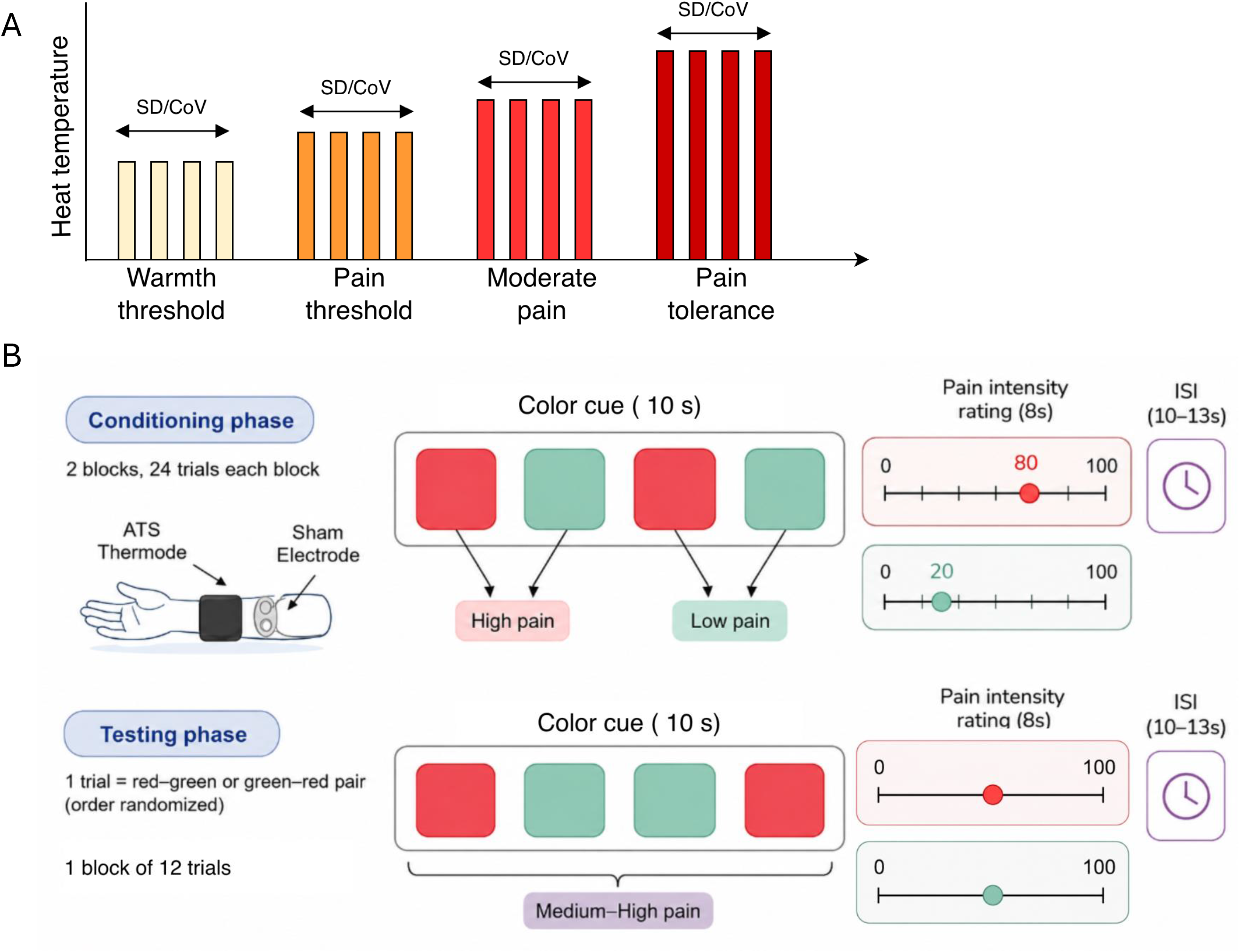
Study design (A). Baseline thermal sensory test. Participants went through warmth threshold, pain threshold, moderate pain, pain tolerance with 4 trials for each modality. Standard deviation (SD) and coefficient of variance (CoV) were calculated for each participant based on the 4 repeated trials for each modality. (B). Classical conditioning procedure to induce placebo effects. Participants completed a 24-trial conditioning phase where a red color cue was paired with high pain stimulation, and a green cue paired with low pain intensity stimulation. Without informing the participants, heat pain stimulation was switched to be identical for both red and green trials during the testing phase (12 trials). The average differences between red and green trials during the testing phase were quantified as the magnitude of placebo effects.

### Conditioning phase

Following calibration, participants completed a conditioning phase consisting of 24 trials. The experimental placebo paradigm was adapted from Colloca and Benedetti 2006 [11] and has been described previously in other publications [8]. Participants were instructed that a sham electrode positioned adjacent to the thermode would deliver an imperceptible electrical stimulation that would reduce pain whenever a green cue appeared on the screen, whereas a red cue indicated that the stimulation was off. Unknown to participants, no electrical stimulation was delivered. Instead, individualized low-painful stimuli (rated approximately 2/10), as measured during the heat pain calibration, were paired with the green cues, and the individualized painful temperature (rated approximately 8/10) was paired with the red cues to establish expectations of analgesia. Importantly, these repeated ratings were repeated 24 times (12 red and12 green) allowing the assessment of pain reliability to comparable pain stimuli.

### Placebo induction

Participants subsequently completed a 12-trial testing phase, during which identical thermal stimuli (the individualized moderate painful temperature) were delivered following both red and green cues (Fig. 2B). Placebo analgesia was quantified as the difference in pain intensity ratings between red-cue (control) and green-cue (placebo) trials, with larger differences indicating greater placebo analgesia. A 100-point Visual Analogue Scale using the Celeritas Response System (Psychology Software Tools Inc., Sharpsburg, PA, USA) was used. Participants showing a reduction in pain rating of 30 points or greater on the Visual Analogue Scale were defined as placebo responders [15].

### Pain-relief expectations

Pain-relief expectations were also assessed using a 100-point VAS ranging from 0=no expectation of pain relief at all to 100=maximum expectation of pain relief. Expectations were measured at the baseline and after the conditioning (e.g., reinforcement expectations).

### Experimenter effect

Given the large number of participants enrolled in the study, we recorded the identity of the experimenter who conducted the calibration and placebo induction procedures for each participant. A total of 16 experimenters administered these procedures, all of whom were trained using the same standardized manual of procedures. To account for potential experimenter effects while ensuring sufficient sample sizes for analysis, experimenters were grouped according to the number of experimental sessions they conducted. An experimental session was defined as the completion of the calibration, conditioning, and testing phases. Three experimenters each completed at least 100 sessions and were retained as individual groups, whereas the remaining 13 experimenters, each of whom conducted fewer than 100 sessions, were combined into a fourth group.

### Statistical Analyses

#### Quantification of pain-reporting variability and reliability

Analyses were designed to characterize individual differences in pain reporting and determine whether these reporter characteristics predicted placebo analgesia. Variability and reliability measures were calculated from repeated heat temperatures obtained during the thermal calibration and placebo conditioning phases controlling for age, sex, race and experimenters.

During the calibration phase, participants received repeated thermal stimuli at predefined temperatures (°C degree), with each stimulus intensity presented four times as described above. These repeated measurements expressed in °C degree were used to quantify individual differences in pain variability and consistency under controlled sensory stimulation. Variability was quantified using the intra-individual standard deviation (SD) and coefficient of variation (CoV). SD reflects the absolute dispersion of repeated pain ratings around an individual’s mean response, whereas CoV represents variability relative to the mean and provides a unitless measure of relative dispersion, allowing comparison across stimulus intensities with different mean pain ratings [1].

During the conditioning phase, repeated pain ratings collected across hypoalgesic-associated and control trials were used to characterize variability in pain reporting during the acquisition of placebo expectations. SD and CoV were calculated separately for each conditioning condition (24 trials). These measures were used to examine whether instability in pain reporting during repeated learning experiences was associated with subsequent placebo analgesia.

#### Pain rating reliability and independent contribution of reporter consistency

Reliability of pain reporting was quantified using the intraclass correlation coefficient (ICC), which estimates the proportion of variance in repeated pain ratings attributable to consistent differences between participants relative to within-participant trial-to-trial variability. Higher ICC values indicate greater consistency of pain ratings across repeated trials, whereas lower ICC values indicate greater inconsistency across assessments. Because ICC and variability indices are derived from the same repeated-measure variance structure, we examined whether reliability provided predictive information beyond conventional variability measures. To determine the unique contribution of reliability, we examined the effects of ICC while controlling for SD and/or CoV.

Prior to analysis, the distribution of continuous variables was evaluated using histograms and Q– Q plots. The variables were judged to be approximately normally distributed, supporting the use of parametric statistics, including Pearson correlation coefficients. Pearson correlations were used to assess associations among variability and reliability measures (SD, CoV, and ICC). Correlation matrices were examined, and associations exceeding |r| ≥ 0.7 were considered indicative of strong collinearity. Variance inflation factors (VIFs) were calculated to evaluate multicollinearity among predictors, with VIF values <5 considered acceptable.

#### Conditioning learning rate

To quantify the acquisition of pain-relief learning during the conditioning phase, we calculated the slope of the red-minus-green pain-rating difference across the 12 conditioning trials using individual-level linear regression models. The standardized regression coefficient was used as each participant’s conditioning learning rate, with larger values indicating stronger learning over time.

#### Placebo analgesia

The magnitude of placebo analgesia was calculated during the testing phase as the difference between pain ratings obtained during control trials (red cue) and placebo trials (green cue). Larger differences indicated greater placebo analgesic responses. Regression models were used to examine whether pain-reporting variability and reliability measures predicted placebo analgesia while accounting for shared variance among reporter characteristics. Analyses focused on quantifying the relationship between conditioning variability and the magnitude of the placebo response while accounting for shared variance between variability measures.

#### Prediction of placebo analgesia

Bivariate correlations (for continuous variables) and univariate ANOVA/t-test (for categorical variables) were used to determine potential sociodemographic covariates including age, sex, race, and experimenters who conducted the study. Variables significantly associated with placebo effects were included as covariates in the main analyses. Namely, age, sex, race (African-American/Black vs. White vs. Asian), and experimenters were included as covariates.

Multiple linear regression models were used to examine whether pain-reporting variability and reliability measures predicted the magnitude of placebo analgesia, treated as a continuous outcome. Models were constructed to evaluate the contribution of variability measures (SD and CoV) and reliability (ICC) and to determine whether reliability explained additional variance beyond variability measures. Improvements in model fit were evaluated by comparing nested models using analysis of variance (ANOVA).

#### Mediation models

Finally, we implemented mediation models to examine whether conditioning learning rate and reinforced expectations mediated the associations of pain-reporting variability with placebo effects. In each model, SD or CoV was considered as the independent variable (X), conditioning learning rate and reinforced expectations were included as parallel mediators, and placebo effects were entered as the dependent variable (Y). Indirect effects were evaluated using 5,000 bootstrap samples to generate 95% confidence intervals. Mediation model was performed using Hayes’ mediation analysis (model 6).

#### K-means clustering

To examine whether patterns of response variability during the conditioning phase were associated with placebo effects, we performed K-means clustering. Clustering variables included the intra-individual SD of pain ratings during the red and green conditioning trials, the ICC of red and green conditioning pain ratings, and the conditioning learning rate (slope of the difference between red and green trial pain ratings). All clustering variables were standardized (z-scores) prior to analysis. We used SD rather than CoV because the two indices were highly correlated (r = 0.86, *p* < 0.001), indicating substantial overlap in the information they captured.

The optimal number of clusters was determined using the average silhouette method, and the solution with the highest average silhouette width was selected. K-means clustering was performed in R using the kmeans() function with the Hartigan–Wong algorithm. Standardized variability measures were clustered using 100 random starting configurations (nstart = 100) and a maximum of 10 iterations per run (iter.max = 10). The optimal number of clusters was determined by jointly examining the elbow method (within-cluster sum of squares) and the average silhouette width across solutions containing two to ten clusters. A random seed of 123 was set to ensure reproducibility.

To examine differences in placebo analgesia across the identified clusters, univariate ANCOVAs were conducted with diagnostic group (TMD vs. HC) and cluster membership as between-subject factors and age, sex, race, and experimenter as covariates. All statistical analyses were performed in SPSS (version 28) and R (version 4.5.2).

## Results

### Participant Characteristics

A total of 801 participants (401 TMD, 400 healthy controls) were included (Table 1). The TMD group was older than healthy controls (41.3 vs 29.4 years) and included a greater proportion of women (76.3% vs 59.5%).

**Table 1.**
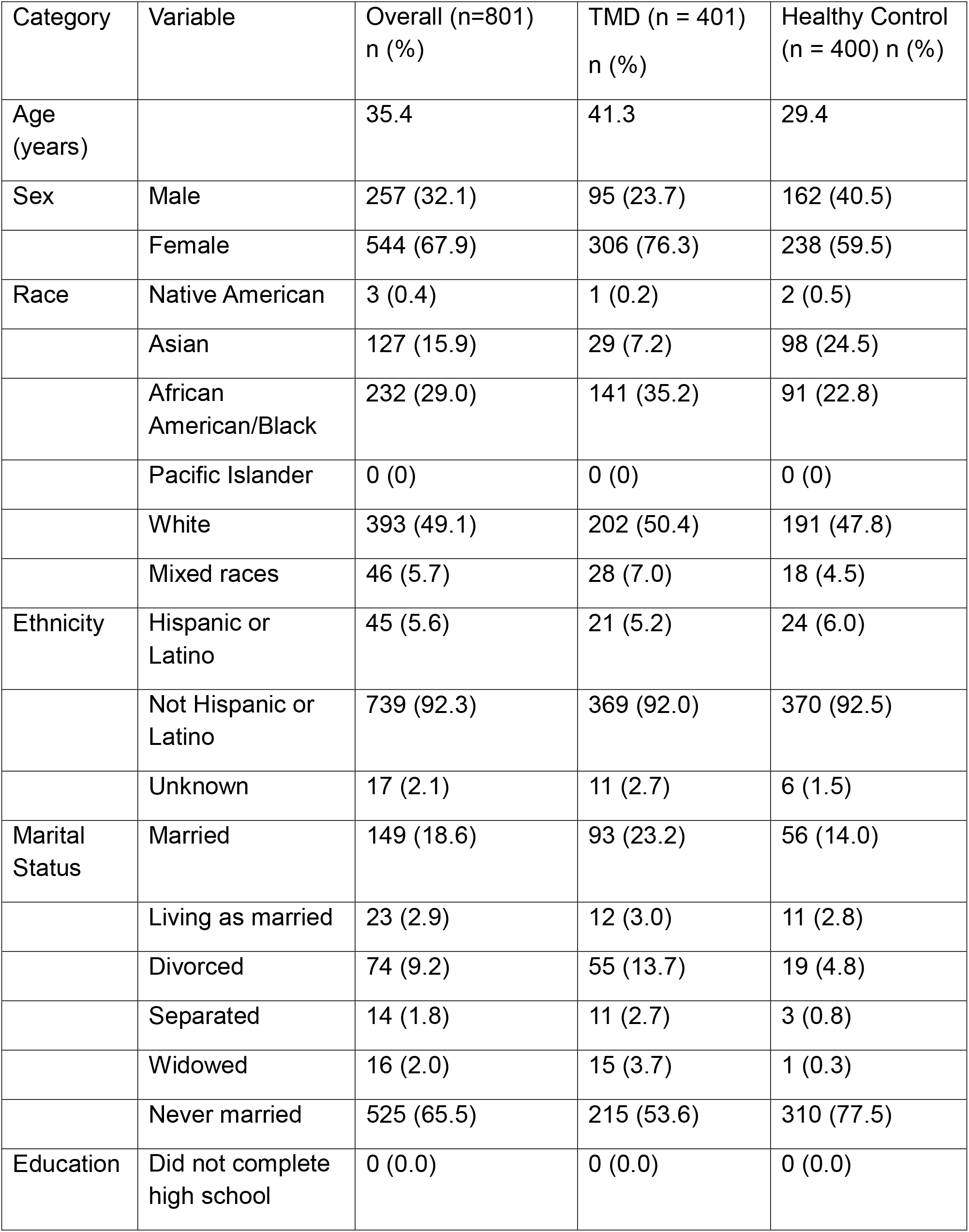
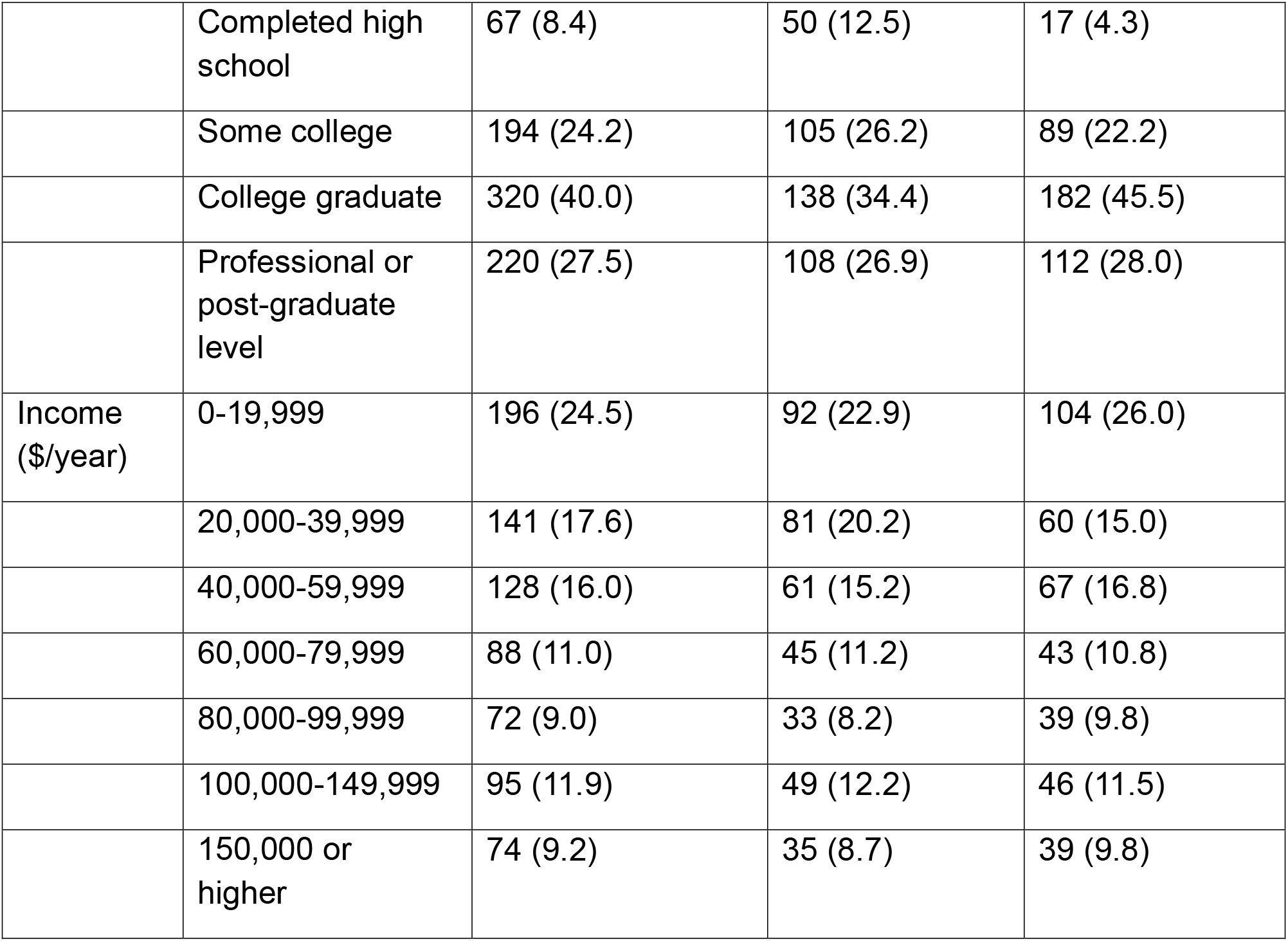
Participant characteristics.

| Category | Variable | Overall (n=801)<br>n (%) | TMD (n = 401)<br>n (%) | Healthy Control<br>(n = 400) n (%) |
| --- | --- | --- | --- | --- |
| Age<br>(years) |  | 35.4 | 41.3 | 29.4 |
| Sex | Male | 257 (32.1) | 95 (23.7) | 162 (40.5) |
|  | Female | 544 (67.9) | 306 (76.3) | 238 (59.5) |
| Race | Native American | 3 (0.4) | 1 (0.2) | 2 (0.5) |
|  | Asian | 127 (15.9) | 29 (7.2) | 98 (24.5) |
|  | African<br>American/Black | 232 (29.0) | 141 (35.2) | 91 (22.8) |
|  | Pacific Islander | 0 (0) | 0 (0) | 0 (0) |
|  | White | 393 (49.1) | 202 (50.4) | 191 (47.8) |
|  | Mixed races | 46 (5.7) | 28 (7.0) | 18 (4.5) |
| Ethnicity | Hispanic or<br>Latino | 45 (5.6) | 21 (5.2) | 24 (6.0) |
|  | Not Hispanic or<br>Latino | 739 (92.3) | 369 (92.0) | 370 (92.5) |
|  | Unknown | 17 (2.1) | 11 (2.7) | 6 (1.5) |
| Marital<br>Status | Married | 149 (18.6) | 93 (23.2) | 56 (14.0) |
|  | Living as married | 23 (2.9) | 12 (3.0) | 11 (2.8) |
|  | Divorced | 74 (9.2) | 55 (13.7) | 19 (4.8) |
|  | Separated | 14 (1.8) | 11 (2.7) | 3 (0.8) |
|  | Widowed | 16 (2.0) | 15 (3.7) | 1 (0.3) |
|  | Never married | 525 (65.5) | 215 (53.6) | 310 (77.5) |
| Education | Did not complete<br>high school | 0 (0.0) | 0 (0.0) | 0 (0.0) |
|  | Completed high school | 67 (8.4) | 50 (12.5) | 17 (4.3) |
|  | Some college | 194 (24.2) | 105 (26.2) | 89 (22.2) |
|  | College graduate | 320 (40.0) | 138 (34.4) | 182 (45.5) |
|  | Professional or post-graduate level | 220 (27.5) | 108 (26.9) | 112 (28.0) |
| Income (\$/year) | 0-19,999 | 196 (24.5) | 92 (22.9) | 104 (26.0) |
|  | 20,000-39,999 | 141 (17.6) | 81 (20.2) | 60 (15.0) |
|  | 40,000-59,999 | 128 (16.0) | 61 (15.2) | 67 (16.8) |
|  | 60,000-79,999 | 88 (11.0) | 45 (11.2) | 43 (10.8) |
|  | 80,000-99,999 | 72 (9.0) | 33 (8.2) | 39 (9.8) |
|  | 100,000-149,999 | 95 (11.9) | 49 (12.2) | 46 (11.5) |
|  | 150,000 or higher | 74 (9.2) | 35 (8.7) | 39 (9.8) |

### Baseline thermal sensory measures and placebo analgesia

During the calibration phase, variability across the four repeated assessments was quantified for each thermal sensory measure (Fig. 3A). Mean temperatures were 34.05°C (range 32.53–44.38) for warmth threshold, 38.61°C (33.07–49.49) for pain threshold, 43.96°C (35.09–51.56) for moderate pain, and 47.88°C (38.95–52.00) for pain tolerance. Mean within-person SDs were 0.53°C, 1.40°C, 1.16°C, and 0.86°C, respectively, whereas mean coefficients of variation (CoV) were 1.49%, 3.58%, 2.66%, and 1.83%.

**Figure 3:**
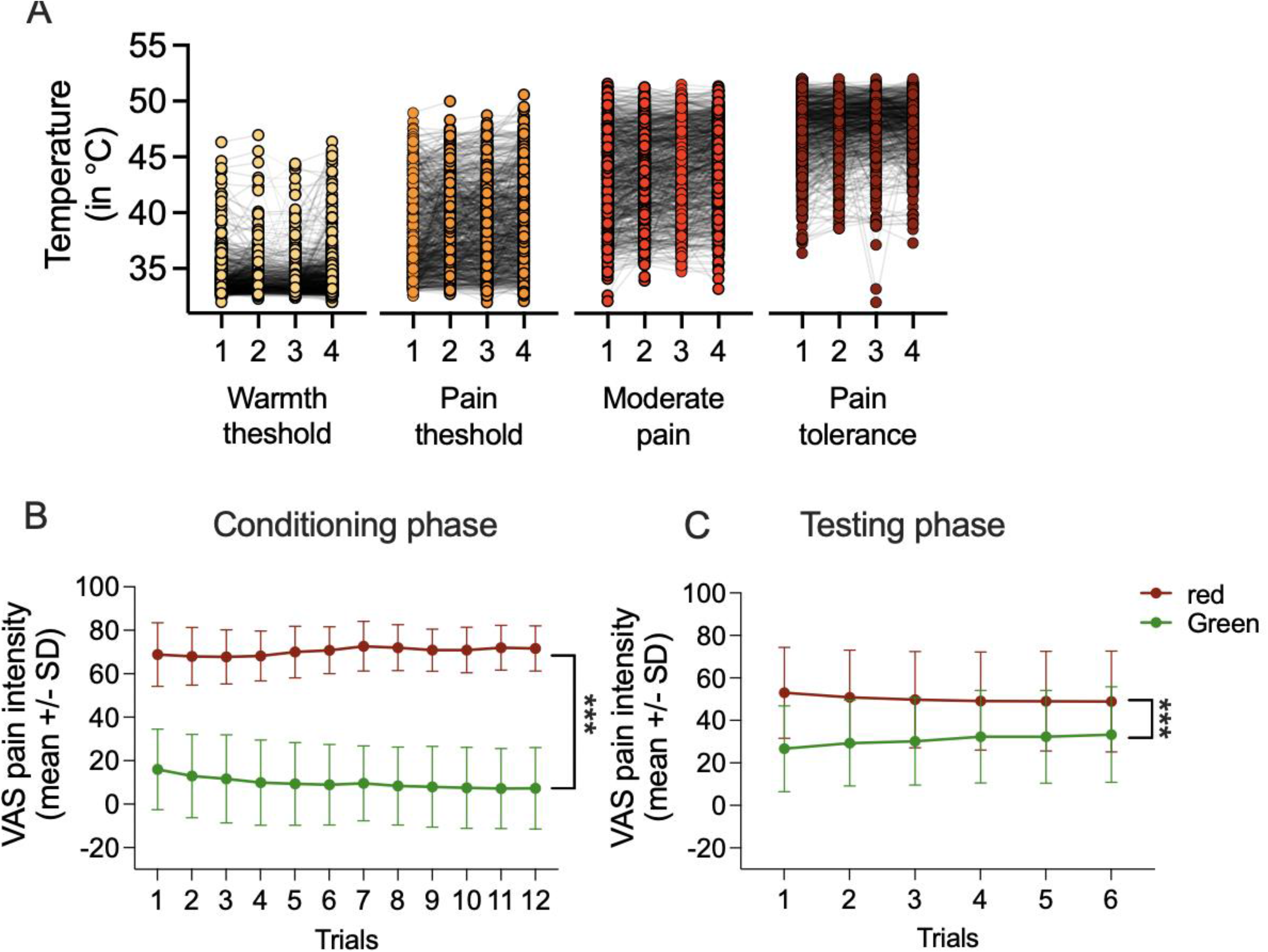
(A) Individual temperature values across four trials for warmth threshold, pain threshold, moderate pain, and pain tolerance. Colored circles represent individual trial values, and gray lines connect repeated measurements from the same participant. (B) Mean visual analog scale (VAS) pain-intensity ratings across 12 conditioning pairs for the red and green cues. Participants reported greater pain intensity corresponding to the red trials as compared to the green trials during the conditioning phase. (C) VAS pain-intensity ratings across six testing pairs for the red and green cues. The pain intensity ratings for the red trials were significantly greater than that in the green trials, suggesting significant placebo effects. Error bars represent standard deviations. Red and green indicate the high-pain and low-pain conditions, respectively. \*\*\**p* < .001.

The conditioning paradigm produced a robust placebo effect. Mean placebo analgesia was 19.4 VAS units, and 60.8% of participants met responder criteria. During the testing phase, pain ratings were significantly lower on placebo (green) than control (red) trials (estimated marginal means: 30.70 ± 0.68 vs. 50.14 ± 0.73 VAS units; *F*(1,796) = 53.40, *p* < .001). The placebo effect did not differ between participants with temporomandibular disorder (TMD) and healthy controls (*F*(1,796) = 0.02, *p* = .889; Fig. 3). Contrary to our hypothesis, variability in baseline thermal sensory measures was not associated with placebo analgesia. Regression models including SD measures explained less than 2% of the variance in placebo response (*R*² = .018), and none of the four thermal measures significantly predicted placebo analgesia (all *p* ≥ .064). Similar findings were obtained using CoV measures (*R*² = .017), with no significant associations observed (all *p* ≥ .074). To determine whether baseline sensory variability influenced the initial expression of placebo analgesia, we next examined placebo effects on the first testing trial. After adjustment for age, sex, race, and experimenter, neither SD nor CoV measures significantly predicted first-trial placebo responses (all *p* ≥ .165).

Likewise, baseline sensory variability was unrelated to placebo effects on the final testing trial, indicating that these measures did not predict placebo responsiveness after repeated testing (all *p* ≥ .310 for SD; all *p* ≥ .322 for CoV).

Among participants with TMD, placebo analgesia did not differ between those with high- and low-impact pain based on the Graded Chronic Pain Scale (*F*(1,312) = 0.06, *p* = .806, see also Table 2). Pain impact also did not moderate the relationship between baseline sensory variability and placebo analgesia for either SD or CoV measures (all *p* > .299).

**Table 2.** Demographic and clinical characteristics of participants with TMD according to pain impact.

| <u>Category</u> | <u>Variable</u> | <u>Overall (n = 401)</u> | <u>Low Impact (n = 227)</u> | <u>High Impact (n = 174)</u> | <u>p low vs high</u> |
| --- | --- | --- | --- | --- | --- |
| <u>Age (years)</u> |  | 41.3 (14.1) | 39.2 (14.3) | 44.2 (13.4) | <0.001 |
| Sex | Male | 95 (23.7) | 58 (25.6) | 37 (21.3) | 0.3 |
|  | Female | 306 (76.3) | 169 (74.4) | 137 (78.7) |  |
| Race | Native American | 1 (0.2) | 1 (0.4) | 0 (0.0) | <0.001 |
|  | Asian | 29 (7.2) | 26 (11.5) | 3 (1.7) |  |
|  | African American/Black | 141 (35.2) | 57 (25.1) | 84 (48.3) |  |
|  | Pacific Islander | 0 (0) | 0 (0) | 0 (0) |  |
|  | White | 202 (50.4) | 134 (59.0) | 68 (39.1) |  |
|  | Mixed races | 28 (7.0) | 9 (4.0) | 19 (10.9) |  |
| Ethnicity | Hispanic or Latino | 21 (5.2) | 10 (4.4) | 11 (6.3) | 0.019 |
|  | Not Hispanic or Latino | 369 (92.0) | 215 (94.7) | 154 (88.5) |  |
|  | Unknown | 11 (2.7) | 2 (0.9) | 9 (5.2) |  |
| Marital Status | Married | 93 (23.2) | 62 (27.3) | 31 (17.8) | 0.019 |
|  | Living as married | 12 (3.0) | 3 (1.3) | 9 (5.2) |  |
|  | Divorced | 55 (13.7) | 27 (11.9) | 28 (16.1) |  |
|  | Separated | 11 (2.7) | 6 (2.6) | 5 (2.9) |  |
|  | Widowed | 15 (3.7) | 5 (2.2) | 10 (5.7) |  |
|  | Never married | 215 (53.6) | 124 (54.6) | 91 (52.3) |  |
| Education | Did not complete high school | 0 (0.0) | <u>0 (0.0)</u> | <u>0 (0.0)</u> | <u>0.002</u> |
|  | Completed high school | 50 (12.5) | <u>19 (8.4)</u> | <u>31 (17.8)</u> |  |
|  | Some college | 105 (26.2) | <u>53 (23.3)</u> | <u>52 (29.9)</u> |  |
|  | College graduate | 138 (34.4) | <u>81 (35.7)</u> | <u>57 (32.8)</u> |  |
|  | Professional or post-graduate level | 108 (26.9) | <u>74 (32.6)</u> | <u>34 (19.5)</u> |  |
| Income (\$/year) | 0-19,999 | 92 (22.9) | <u>36 (16.0)</u> | <u>56 (32.7)</u> | <u><0.001</u> |
|  | 20,000-39,999 | 81 (20.2) | <u>47 (20.9)</u> | <u>34 (19.9)</u> |  |
|  | 40,000-59,999 | 61 (15.2) | <u>29 (12.9)</u> | <u>32 (18.7)</u> |  |
|  | 60,000-79,999 | 45 (11.2) | <u>32 (14.2)</u> | <u>13 (7.6)</u> |  |
|  | 80,000-99,999 | 33 (8.2) | <u>21 (9.3)</u> | <u>12 (7.0)</u> |  |
|  | 100,000-149,999 | 49 (12.2) | <u>32 (14.2)</u> | <u>17 (9.9)</u> |  |
|  | 150,000 or higher | 35 (8.7) | <u>28 (12.4)</u> | <u>7 (4.1)</u> |  |
| Placebo score |  | 18.6 (17.7) | <u>18.5 (16.2)</u> | <u>18.6 (19.6)</u> | 0.5 |
| Placebo Responder | No | 195 (46.1) | <u>98 (43.2)</u> | <u>87 (50.0)</u> | 0.2 |
|  | Yes | 216 (53.9) | <u>129 (56.8)</u> | <u>87 (50.0)</u> |  |

Note: Continuous variables are presented as mean (SD), and categorical variables as *n* (%). Between-group differences were assessed using independent-samples *t*-tests for continuous variables and $\chi^2$ tests for categorical variables.

These findings indicate that variability in baseline thermal sensory responses does not meaningfully predict placebo analgesia, either during the initial expression of placebo responding or after repeated placebo testing.

### Trial-to-trial pain variability during placebo conditioning

Because baseline sensory variability was not associated with placebo analgesia, we next examined whether trial-to-trial variability during placebo conditioning—the phase in which cue– pain associations are acquired—predicted subsequent placebo responses.

Greater variability (SD) in pain ratings during reinforced red trials significantly predicted smaller placebo effects (β = −0.16, *p* < .001; Fig. 4A), whereas variability during green trials was not associated with placebo analgesia (*p* = .118). The association between red-trial variability and placebo analgesia remained significant after adjustment for reinforced expectancy ratings (β = −0.16, *p* < .001), indicating that conditioning variability explained unique variance beyond expectancy. The overall model explained 11% of the variance in placebo response (*R*² = .110). Among the covariates, age (*p* = .011) and experimenter (*p* < .001) were significantly associated with placebo responses, whereas sex (*p* = .108) and participant group (*p* = .579) were not.

**Figure 4.**
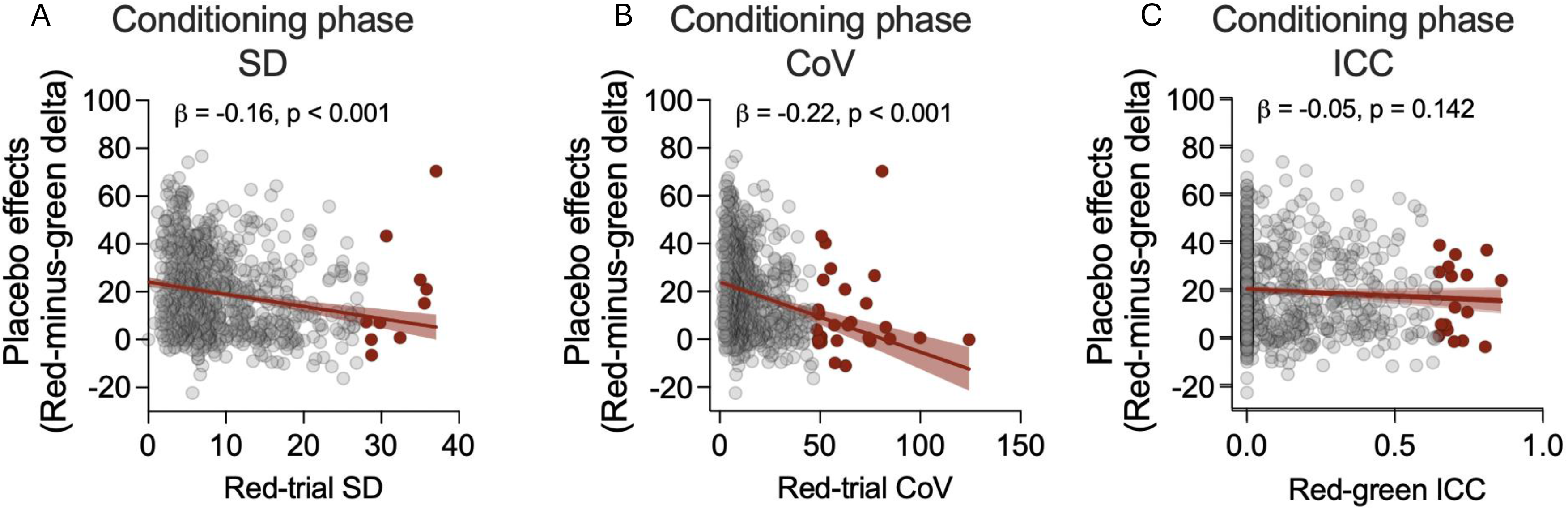
Associations between conditioning-phase variability and placebo effects. Scatterplots show the associations of red-trial standard deviation (A), coefficient of variation (B), and red/green intraclass correlation coefficient (C) with placebo effects, calculated as the averaged difference in pain ratings between the red and green conditions. Solid lines represent fitted regression lines, and shaded areas indicate 95% confidence intervals. Gray circles represent individual participants; red circles highlight outliers. Greater red trial SD and CoV were associated with smaller placebo effects, whereas red/green ICC was not significantly associated with placebo effects. Removing the outliers did not change these findings.

We next examined whether this relationship differed between healthy controls and participants with temporomandibular disorder (TMD). Although the group-by-red SD interaction did not reach statistical significance (*F*(1,787) = 3.61, *p* = .058), stratified analyses indicated that the association was evident in participants with TMD (β = −0.23, *p* < .001) but not in healthy controls (β = −0.07, *p* = .174).

Analyses using the coefficient of variation (CoV) yielded similar results. Greater red-trial CoV predicted smaller placebo effects (β = −0.22, *p* < .001; Fig. 4B), whereas green-trial CoV was not associated with placebo analgesia (*p* = .179). The group-by-red CoV interaction was not significant (*F*(1,760) = 1.89, *p* = .169). These findings remained unchanged after excluding outliers.

Finally, we examined whether baseline sensory variability was associated with variability during placebo conditioning. Baseline sensory variability showed only weak associations with conditioning variability. Greater pain tolerance SD predicted greater variability during both red (β = 0.15, *p* < .001) and green (β = 0.12, *p* = .008) conditioning trials, whereas pain tolerance CoV predicted greater red-trial CoV (β = 0.15, *p* < .001) but not green-trial CoV (*p* = 1.000). Variability in warmth and pain thresholds was not associated with conditioning variability (all *p*s > .596). All *p*-values were Bonferroni-corrected for multiple comparisons.

### Reliability and consistency of pain ratings

We next examined whether the reliability of pain reporting, quantified using the ICC, predicted placebo analgesia. ICC during the conditioning phase was not associated with placebo effects (*F*(1,762) = 0.02, *p* = .894; Fig. 4C). In contrast, higher ICC during the testing phase was associated with smaller placebo effects (β = −0.13, *F*(1,782) = 13.70, *p* < .001), and this association remained significant after adjustment for testing-phase variability (red and green trial SDs; β = −0.12, *p* < .001). The group-by-testing ICC interaction was not significant (*F*(1,782) = 0.45, *p* = .503). However, because testing-phase ICC was derived from the same trials used to calculate placebo analgesia, this association is unlikely to represent a prospective predictor and more likely reflects shared measurement variance.

### K-means clustering of conditioning responses

The silhouette and elbow methods indicated that a three-cluster model provided the best fit to the data. K-means clustering identified three distinct conditioning-response profiles (Fig. 5A). Cluster 1 (*n* = 134) was characterized by high trial-to-trial variability (SD and CoV) during both red and green conditioning trials, intermediate ICC, and intermediate learning rates, representing an unstable learning profile. Cluster 2 (*n* = 465) showed low variability, the lowest ICC, and the highest learning rates, representing a stable, strong-learning profile. Cluster 3 (*n* = 175) also exhibited low variability but had the highest ICC and the lowest learning rates, representing a stable but weak-learning profile. Cluster membership differed by TMD status (χ²(2) = 21.43, *p* < .001). Participants with TMD were more likely to belong to the unstable learning cluster (23.4% vs. 11.0%), whereas healthy controls were more likely to belong to the stable, strong-learning cluster (65.9% vs. 54.5%). Cluster membership also differed by race (χ²(6) = 61.53, *p* < .001), but not by sex (χ²(2) = 0.95, *p* = .621). Participants in the unstable learning cluster were older than those in the other two clusters (*F*(2,771) = 12.43, *p* < .001). In addition, this cluster exhibited greater baseline pain-tolerance variability (SD and CoV) than either stable learning cluster (all Bonferroni-adjusted *p*s < .01).

**Figure 5.**
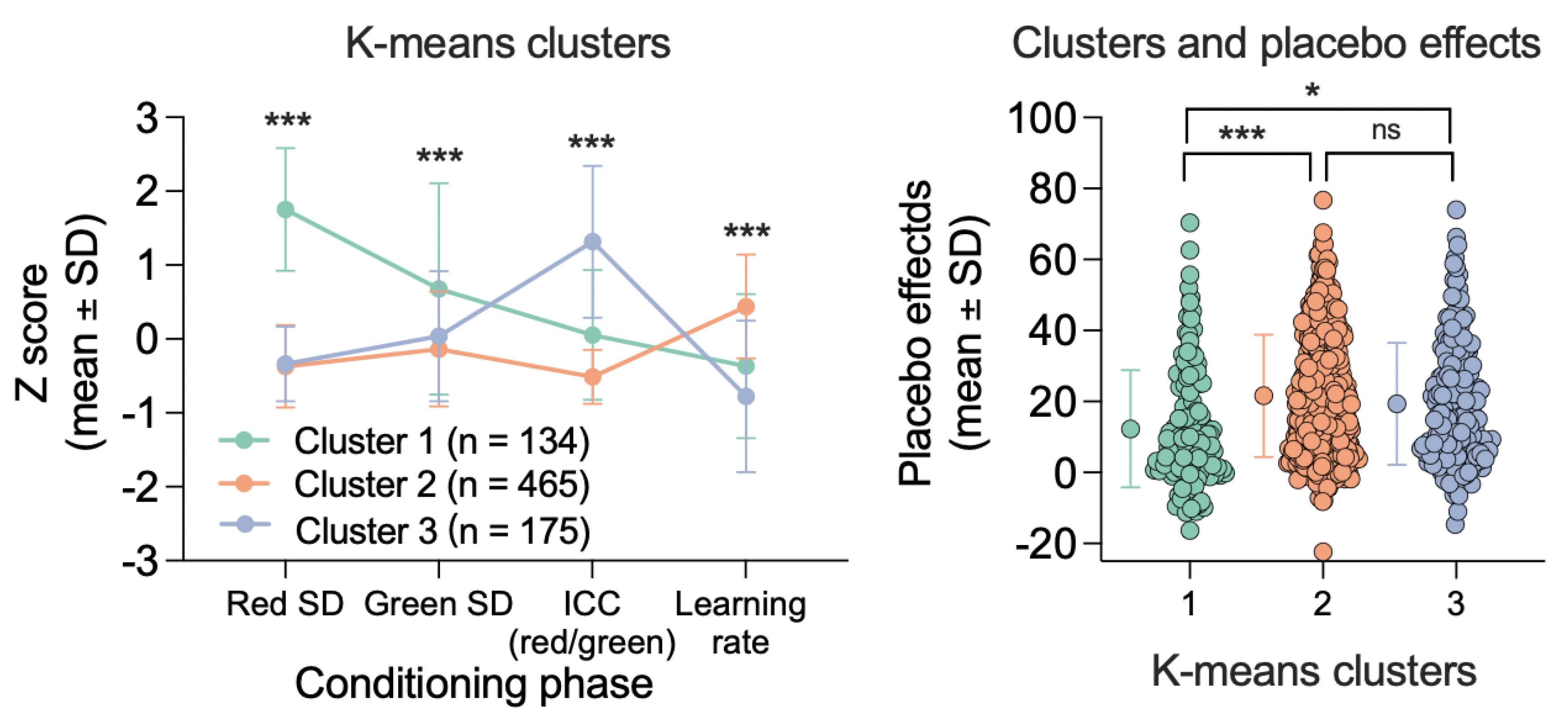
K-means clustering of conditioning profiles and cluster differences in placebo effects. **(A)** Standardized mean scores for red-trial SD, green-trial SD, red/green intraclass correlation coefficient (ICC), and learning rate across the three K-means clusters. Error bars represent standard deviations. Cluster 1 was characterized by high trial-to-trial variability during the conditioning phase and medium learning rate, Cluster 2 by low variability and stronger learning, and Cluster 3 by relatively stable responses but weaker learning. The right panel shows placebo effects during the testing phase for each cluster. Points represent individual participants, and error bars represent means ± SD. Cluster 1 showed significantly smaller placebo effects than Clusters 2 and 3, whereas Clusters 2 and 3 did not differ significantly. * *p* < .05; \*\*\**p* < .001; ns, not significant.

### Learning profiles and placebo analgesia

We next examined whether the identified learning profiles differed in placebo analgesia. After adjustment for age, sex, race, and experimenter, cluster membership was significantly associated with placebo response (*F*(2,764) = 9.42, *p* < .001). Participants in the unstable learning cluster exhibited significantly smaller placebo effects (mean = 13.81, SEM = 1.58) than those in the stable, strong-learning cluster (mean = 21.48, SEM = 0.78, *p* < .001) and the stable, weak-learning cluster (mean = 19.15, SEM = 1.26, *p* = .026; Fig. 5B). Placebo analgesia did not differ between the two stable learning clusters (*p* = .354), suggesting that learning stability, rather than the rate or magnitude of conditioning, was the primary determinant of placebo responsiveness. The association between learning profiles and placebo analgesia did not differ between participants with TMD and healthy controls (group × cluster interaction: *F*(2,764) = 1.87, *p* = .154).

### The Experimenter effects

Experimenter identity was associated with several study measures, including variability in baseline thermal pain sensitivity (pain threshold and pain tolerance), trial-to-trial variability during placebo conditioning (red and green trials), and the magnitude of placebo analgesia (all *p*s < .01). In contrast, experimenter identity was not associated with the conditioning learning rate (Red-minus-Green slope; *F*(3,796) = 1.94, *p* = .122). Given these effects, experimenter identity was included as a covariate in all primary regression and mixed-effects models. Importantly, the associations between conditioning-phase pain-report variability and placebo analgesia remained significant after adjustment for experimenter, indicating that the findings were not attributable to experimenter-related differences.

### Mediation by conditioning learning and expectancy

To examine potential mechanisms, we tested whether conditioning learning rate and post-conditioning expectancy mediated the association between conditioning variability and placebo analgesia (Fig. 6). For both SD and CoV models, greater variability during reinforced (red) conditioning trials was associated with slower conditioning learning (*p*s < .001), which in turn predicted smaller placebo effects (*p*s < .001). Although greater variability was also associated with lower post-conditioning expectancy (*p*s < .05), expectancy was not independently associated with placebo analgesia (*r* = 0.01, *p* = .833; all mediation paths *p*s > .58). These findings suggest that conditioning variability reduces placebo analgesia primarily through impaired learning rather than altered expectancy.

**Figure 6.**
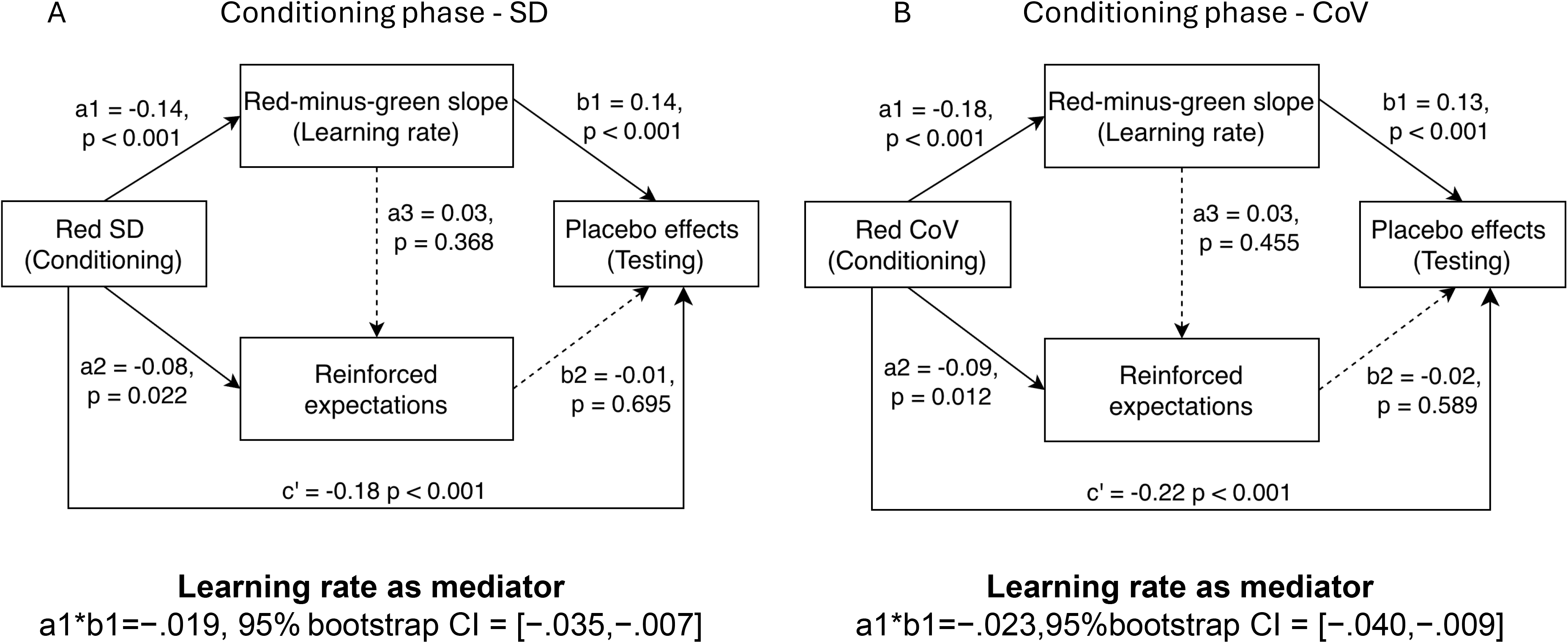
Mediation models. The models examined whether learning rate and reinforced expectations sequentially mediated the associations of red-trial standard deviation (SD; A) and coefficient of variation (CoV; B) with placebo effects during testing. Solid arrows indicate statistically significant paths, whereas dashed arrows indicate nonsignificant paths. Greater red-trial SD and CoV were associated with weaker learning rates and lower reinforced expectations. Stronger learning rates predicted larger placebo effects; however, reinforced expectations were not independently associated with placebo effects. Significant direct effects of red-trial SD and CoV on placebo effects remained after accounting for the mediators. Values are standardized regression coefficients with corresponding *p* values.

## Discussion

Variability in baseline sensory measures, including warmth threshold, pain threshold, moderate pain, and pain tolerance, showed no associations with placebo analgesia. We identified trial-to-trial pain variability during placebo conditioning as a predictor of subsequent placebo analgesia. Participants who exhibited greater instability in conditioned pain ratings, particularly during reinforced high-pain (red) trials, showed smaller placebo effects during testing. This association was specific to the conditioning phase and was not explained by baseline sensory variability, indicating that general fluctuations in pain perception do not account for individual differences in placebo responsiveness. Instead, variability during conditioning appears to reflect differences in the acquisition of cue–pain associations. Consistent with this interpretation, conditioning learning rate mediated the relationship between pain-report variability and placebo analgesia, whereas post-conditioning reinforced expectations did not. These findings suggest that the stability with which individuals encode and update pain experiences during learning, rather than baseline sensory variability or explicit expectancy alone, is an important determinant of placebo responsiveness. Whether conditioning-phase pain variability has predictive value in clinical treatment contexts remains to be established.

A growing body of work has examined whether within-subject variability in pain reports can serve as a marker of placebo responsiveness [35] postulating that within-subject variability could be viewed as a proxy measure for the precision/certainty in ascending sensory signals, which in turn could predict placebo analgesia [23; 32]. However, findings have been inconsistent, possibly because variability has typically been assessed before treatment exposure and without considering the phase of placebo learning. In a crossover trial of naproxen versus placebo in patients with knee osteoarthritis (n=55), greater variability in clinical pain ratings and experimental pain responses measured with the Focused Analgesia Selection Test (FAST) was associated with placebo responsiveness (r=0.393, P=0.004 and r=−0.371, P=0.009, respectively) [36]. FAST variability predicted the treatment difference between naproxen and placebo, suggesting that experimentally induced pain variability may capture individual differences relevant to treatment responsiveness. However, subsequent studies have questioned the generalizability of baseline pain variability as a predictive marker [34]. Analyses from crossover trials in peripheral neuropathic pain found that baseline pain coefficient of variation, standard deviation, and range did not significantly associated with the placebo response [19]. In our study, baseline sensory variability showed minimal association with placebo responsiveness.

Variability measured during experimental pain may reflect mechanisms related to associative learning and expectancy rather than spontaneous fluctuations in clinical pain. The absence of meaningful associations between thermal (heat) sensory quantitative test variability and placebo effects may also provide insight into the mechanisms underlying placebo analgesia. Contemporary models emphasize expectancy, learning, motivation, and endogenous pain-modulatory systems as central drivers of placebo effects [3; 5; 6; 9; 12; 18; 27; 40]. If placebo responsiveness were primarily determined by instability in sensory processing [23], stronger relationships would be expected between variability in warmth detection, pain threshold, or pain tolerance and subsequent (conditioned) placebo effects. Instead, the weak associations observed here suggest that placebo analgesia depends more heavily on higher-order cognitive and learning processes [7; 11; 14] than on variability at the level of sensory detection.

The critical role of the learning process was further supported by mediation analyses showing that the slope of red–green pain rating differences during the conditioning phase (i.e., the rate of acquisition of the placebo contingency) significantly mediated the association between conditioning-phase variability (con_red_SD) and placebo effects. Specifically, greater trial-to-trial variability during conditioning was associated with a slower rate of learning, which in turn was associated with a reduced placebo effect. This finding suggests that the detrimental effect of higher variability is not attributable to a generalized tendency to provide unreliable pain ratings but rather reflects differences in how effectively individuals acquire and update expectations based on repeated cue–outcome experiences. In other words, greater variability during the learning phase may interfere with the formation of a stable predictive relationship between the treatment cue and pain relief, ultimately weakening subsequent placebo responses. These findings may also have implications for the design and interpretation of experimental analgesic studies. Participants exhibiting greater pain-reporting variability during conditioning developed smaller placebo effects, suggesting that instability during learning may reduce the consistency with which contextual cues modulate pain. If this instability also increases trial-to-trial variability in placebo analgesia, it could increase variability in the placebo-adjusted treatment effect (i.e., the difference between active treatment and placebo conditions), thereby reducing statistical power to detect true analgesic effects.

Reliability measures showed stronger associations with placebo response than traditional variability metrics [24]. Models incorporating intraclass correlation coefficients explained additional variance beyond measures of dispersion, suggesting that response consistency may capture aspects of placebo responsiveness not reflected by standard variability indices. However, these associations should be interpreted cautiously because testing-phase reliability was calculated from the same observations used to derive placebo response, resulting in substantial predictor–outcome overlap and potential mathematical coupling. In contrast, conditioning-phase reliability remained associated with placebo response after residualization, although the effect sizes were modest. These findings suggest that response reliability may contribute to individual differences in placebo analgesia, but its predictive value is limited and insufficient for identifying placebo responders at the individual level.

One limitation is the generalizability of these findings needs to be considered. We used a conditioning placebo induction paradigm (see Clusters based on learning slope), and socially or verbally [29] driven placebo effects may respond differently. Also, these results may not necessarily translate to placebo arms of randomized clinical trials. Additionally, regression to the mean has been proposed as a potential contributor to apparent placebo responses in clinical trials, particularly when participants are selected based on high baseline pain levels [22]. However, because baseline thermal pain variability was unrelated to placebo analgesia and our analyses focused on experimentally induced placebo responses following conditioning, regression to the mean is unlikely to account for the observed associations. Moreover, despite the experimenters (four) being trained in a standardized way to conduct the calibration, conditioning, and placebo induction, we observed a significant experimenter effect on the calibration, conditioning and placebo phases. The result was driven by one experimenter who may have excelled in the ability to engage and connect with study participants across groups. Using empathy scales for doctor-patient interactions [21] may help control and quantify for the effects of the experimenter on placebo magnitudes.

Several strengths of this study should also be highlighted. First, the large and well-characterized sample, including both individuals with TMD and healthy controls, provided sufficient statistical power to detect small effects and allowed a rigorous evaluation of multiple candidate predictors of placebo responsiveness. Second, the experimental conditioning paradigm enabled the investigation of variability and reliability during the acquisition of placebo learning, prior to placebo response assessment, providing a stronger temporal framework for evaluating potential mechanisms than would be possible in purely observational studies. Third, the use of repeated trial-level pain ratings allowed examination of dynamic features of pain processing, including variability, reliability, and learning trajectories, rather than relying solely on single summary measures of sensory sensitivity. Finally, the combination of regression, residualized analyses, mediation models, and classification approaches provided a comprehensive assessment of whether pain-reporting characteristics could meaningfully explain or predict individual differences in placebo analgesia.

Experimental pain paradigms may provide a useful bridge between laboratory and clinical research. Because experimentally evoked pain can be assessed before therapeutic interventions, future studies should determine whether variability during standardized pain testing predicts placebo responsiveness or treatment outcomes in acute clinical pain settings, where pre-treatment clinical pain variability cannot be readily quantified.

## Conclusion

Variability in baseline sensory measures, including warmth threshold, pain threshold, moderate pain, and pain tolerance provided limited value for identifying placebo responders. In contrast, trial-to-trial variability during placebo conditioning, particularly during reinforced high-pain (red) trials, emerged as a prospective predictor of placebo responsiveness. Greater instability in conditioned pain responses was associated with smaller placebo effects, with consistent findings across both SD and CoV measures. Mediation analyses indicated that this association was primarily linked to reduced conditioning learning rate, whereas post-conditioning expectancy did not account for the relationship. Together, these findings suggest that pain-report variability is not a general characteristic of sensory processing but rather a phase-specific marker of the ability to acquire and maintain stable placebo-related associations. These findings provide a mechanistic framework for understanding heterogeneity in placebo analgesia by showing that variability during placebo learning, rather than baseline sensory variability, is associated with subsequent placebo responsiveness.

## Data Availability

All data produced in the present study are available upon reasonable request to the authors

## Acknowledgements

This research is supported by National Institute Dental Craniofacial Research, NIDCR (R01 DE025946, LC and R21 DE032532 LC/SGD). The funding agencies have no role in the study. Participant compensation was financed with funds from (R01 DE025946, LC. 35% of the efforts of L.C. and SGD, 100% of YW, were financed with federal funds. The content is solely the responsibility of the authors and does not necessarily represent the official views of the National Institutes of Health.

